# Reassessing the management of uncomplicated urinary tract infection: A retrospective analysis using machine learning causal inference

**DOI:** 10.1101/2024.08.18.24312104

**Authors:** Noah C Jones, Ming-Chieh Shih, Elizabeth Healey, Chen Wen Zhai, Sonali D Advani, Aaron Smith-McLallen, David Sontag, Sanjat Kanjilal

**Author notes:** **CORRESPONDING AUTHOR** Sanjat Kanjilal MD, MPH, Department of Population Medicine, Harvard Medical School & Harvard Pilgrim Healthcare Institute, 401 Park Drive, Suite 401 East, Boston, MA 02215. Co-last author (David Sontag, Sanjat Kanjilal).

## Abstract

**Background:** Uncomplicated urinary tract infection (UTI) is a common indication for outpatient antimicrobial therapy. National guidelines for the management of uncomplicated UTI were published by the Infectious Diseases Society of America in 2011, however it is not fully known the extent to which they align with current practices, patient diversity, and pathogen biology, all of which have evolved significantly in the time since their publication.

**Objective:** We aimed to re-evaluate efficacy and adverse events for first-line antibiotics (nitrofurantoin, and trimethoprim-sulfamethoxazole), versus second-line antibiotics (fluoroquinolones) and versus alternative agents (oral β-lactams) for uncomplicated UTI in contemporary clinical practice by applying machine learning algorithms to a large claims database formatted into the Observational Medical Outcomes Partnership (OMOP) common data model.

**Outcomes:** Our primary outcome was a composite endpoint for treatment failure, defined as outpatient or inpatient re-visit within 30 days for UTI, pyelonephritis or sepsis. Secondary outcomes were the risk of 4 common antibiotic-associated adverse events: gastrointestinal symptoms, rash, kidney injury and *C. difficile* infection.

**Statistical methods:** We adjusted for covariate-dependent censoring and treatment indication using a broad set of domain-expert derived features. Sensitivity analyses were conducted using `OMOP-learn`, an automated feature engineering package for OMOP datasets.

**Results:** Our study included 57,585 episodes of UTI from 49,037 patients. First-line antibiotics were prescribed in 35,018 (61%) episodes, second-line antibiotics were prescribed in 21,140 (37%) episodes and alternative antibiotics were prescribed in 1,427 (2%) episodes. After adjustment, patients receiving first-line therapies had an absolute risk difference of -2.1% [95% CI -2.9% to -1.6%] for having a revisit for UTI within 30 days of diagnosis relative to second-line antibiotics. First-line therapies had an absolute risk difference of -6.6% [95% CI -9.4% to -3.8%] for 30-day revisit compared to alternative β-lactam antibiotics. Differences in adverse events were clinically similar between first and second line agents, but lower for first-line agents relative to alternative antibiotics (−3.5% [95% CI -5.9% to -1.2%]). Results were similar for models built with `OMOP-learn`.

**Conclusion:** Our study provides support for the continued use of first-line antibiotics for the management of uncomplicated UTI. Our results also provide proof-of-principle that automated feature extraction methods for OMOP formatted data can emulate manually curated models, thereby promoting reproducibility and generalizability.

## INTRODUCTION

Up to 50% of women will experience a urinary tract infection (UTI) in their lifetime^1^, making it the third most common indication for antibiotic treatment in the United States after respiratory tract infection and skin and soft tissue infections^2^. Treatment guidelines published by the Infectious Diseases Society of America (IDSA) encourage the use of nitrofurantoin, trimethoprim-sulfamethoxazole and fosfomycin as first-line treatments for uncomplicated UTI based on their efficacy and relatively limited side effect profile^3,4^. Fluoroquinolones are listed as a second-line option due to their predilection for selecting for multidrug resistant organisms^5^ and their association with serious adverse events including *C. difficile* colitis^6^. Despite this, ciprofloxacin and levofloxacin are still the most commonly used antibiotics in the treatment of UTI, which may reflect the real or perceived threat of antibiotic resistance to the first line agents^2^. The guidelines list beta-lactams as alternative treatments as they are associated with reduced treatment efficacy^7^.

The evidence base supporting the IDSA treatment guidelines are based on a small number of randomized controlled trials and observational studies ^7–11^, many of which were completed several decades ago. While these provide important information for policy making, they were limited in the diversity of patients they recruited and were performed at a time when standards of care, and health-seeking behavior differed significantly from current practice. Furthermore, the pathogen strains in circulation at the time of these studies have likely been replaced by new strains that may have a differential response to drug therapies regardless of their susceptibility phenotype. Therefore, a re-evaluation of management strategies for uncomplicated UTI could provide useful information for treating clinicians and policy makers. In this study, we sought to estimate treatment efficacy and adverse events for guideline-concordant and discordant treatments for UTI using causal inference supported by machine learning applied to a large contemporary claims dataset.

## METHODS

### Study design and data

We conducted a retrospective cohort study using the claims database from Independence Blue Cross, which contains health-related information for 3 million people living primarily in a 5 county area surrounding Philadelphia, PA. The dataset contains inpatient, outpatient, laboratory and pharmacy claims made between 2012 and 2021. The database is formatted in the Observational Medical Outcomes Partnership (OMOP) common data model (version 5), developed by the Observational Health Data Sciences and Informatics (OHDSI) initiative^12^. Reporting of this study follows the Strengthening the Reporting of Observational Studies in Epidemiology (STROBE) statement^13^. This study was deemed exempt by the Institutional Review Board of the Massachusetts Institute of Technology (protocol E-3970).

### Study population

The analysis cohort consisted of non-pregnant females aged 18 and older with a diagnosis of uncomplicated, non-recurrent UTI at an outpatient setting. The list of diagnosis codes associated with a diagnosis of UTI is provided in Supplementary Table 1. Patient included in the analysis must also have been treated with one of the following three classes antimicrobials within a 7-day period after the diagnosis: a) nitrofurantoin, and trimethoprim-sulfamethoxazole (first line treatments), b) ofloxacin, ciprofloxacin and levofloxacin (second-line treatments), or c) specific oral β-lactam drugs used for UTI such as amoxicillin-clavulanate, cefadroxil, and cefpodoxime (alternative treatments). Fosfomycin was excluded due to the low number of treatment events.

We excluded individuals with UTI who received treatment outside of the three classes above, e.g. fluconazole, and individuals treated with more than one antibiotic within a 7-day period. In addition, to avoid contamination of previous antibiotic exposures, we excluded patients that had antibiotic exposure within 7 days before the date of UTI diagnosis. We also excluded those with recurrent UTI, defined as ≥2 episodes in a 180 day period and ≥3 episodes in a 365 day period, and people with complicated UTI, defined as any males with a UTI diagnosis or females with a predefined list of procedures and diagnoses associated with complicated UTI within 180 days of the diagnosis, or any histories of complicating long-term comorbidities such as neurogenic bladder, spina bifida, or cancers of the genitourinary tract prior to the UTI diagnosis. A full list of comorbidities flagged for exclusion can be found in Supplementary Table 2.

### Outcomes and Censoring

We defined two primary endpoints for the analysis. The first was a composite endpoint for treatment failure, defined as outpatient or inpatient re-visit within 30 days for UTI, pyelonephritis or sepsis. The second set of endpoints involved adverse events, defined as the presence of diarrhea within 15 days of the UTI event, acute kidney injury (AKI) or a dermatologic adverse event within 30 days of a UTI event or a diagnosis of *C. difficile* infection within 90 days of the UTI event. The conditions and the corresponding codes included in each adverse event category are listed in **Error! Reference source not found**.. Individuals were right-censored from the analysis if they left the health plan before the observational period of the outcome of interest.

### Confounder generation

We derived 2 sets of baseline covariates, which served as potential confounders. The first utilized domain expert knowledge from two practicing infectious disease physicians (SA and SK), and the second was derived from the `OMOP-learn` coding package. `OMOP-learn` is a data-driven feature extractor developed in prior work and is specifically designed for use with claims datasets formatted in the OMOP common data model^14^. The package serves to automatically generate time-windowed covariates.

Domain expert-derived features were classified into demographics, medical conditions, drug prescriptions, prior UTI history, prior antibiotic exposures, laboratory measurements, and provider specialty and year of prescription to account for secular trends in prescribing behavior. A list of medical conditions and drug prescriptions included in the features is shown in Supplementary Table 4. Domain expert-derived features related to medical histories were binned into non-overlapping time windows of 0 to ≤6 months, >6 to ≤12 months and >12 to ≤24 months relative to the date of UTI diagnosis. Laboratories (urinalysis and blood tests) were counted if they were drawn at the time of the UTI diagnosis. The final expert-derived model consisted of 245 features. `OMOP-learn` features were derived from diagnoses, procedures, medications, and provider specialties. The total OMOP-derived model consisted of 143,830 features for the comparison between first and second-line antibiotics, and 131,035 features for the comparison between first-line and alternative antibiotics. Figure 1 depicts the cohort, outcome and confounder definitions.

**Figure 1:**
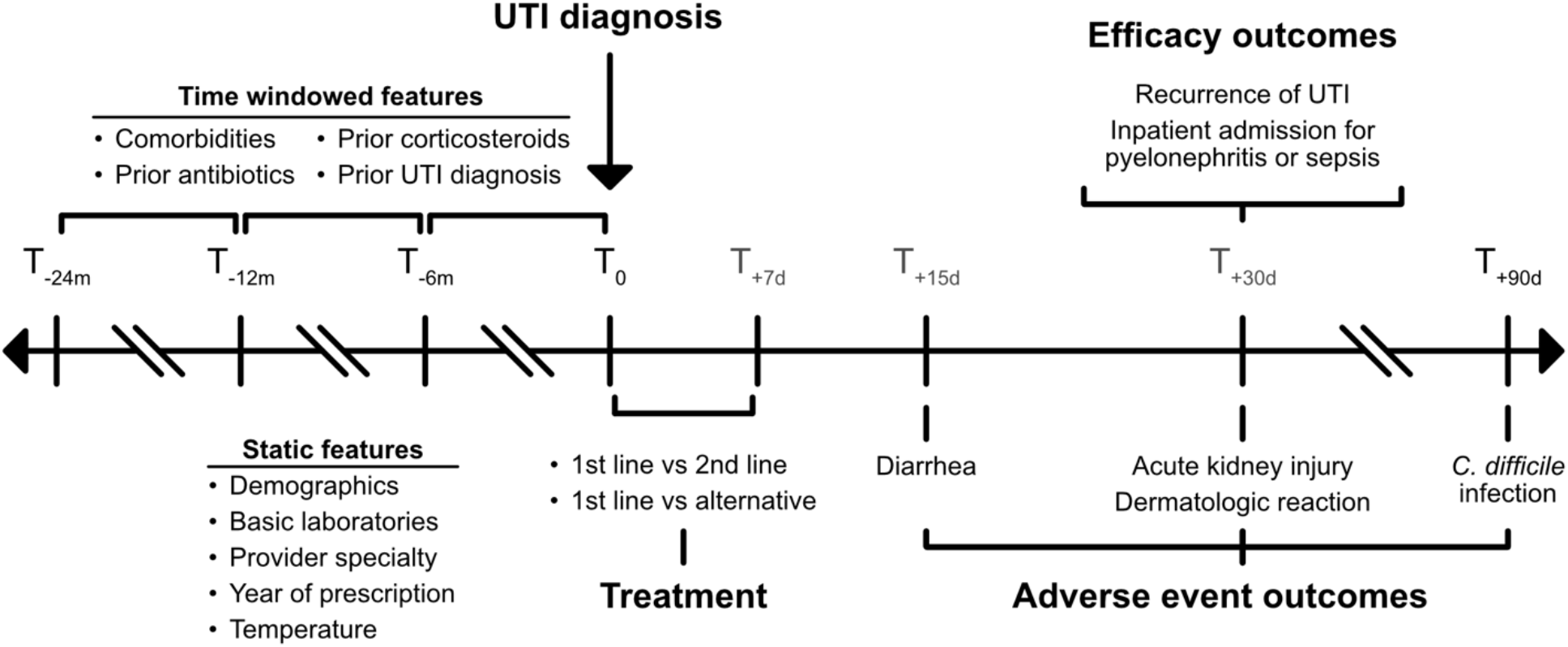
Cohort inclusion criteria and definitions for outcomes and feature. Static features were evaluated at T_0_. Abbreviations, T, time.

### Statistical analysis

We estimated the absolute risk difference of first line therapies versus second line or alternative therapies for patients with uncomplicated UTI on 30-day recurrence and adverse effects. To account for possible covariate-dependent censoring, we used inverse probability of censoring weighting (IPCW) to reweight individuals that were observed or not censored^15^. In addition, the central problem in estimating antibiotic treatment on outcomes is confounding by indication, therefore we utilized inverse probability weighted propensity scores to adjust for the likelihood of receipt of each treatment class given an individual’s confounders.

For both the observation probability model and treatment propensity score model, the dataset was split 80/20 into a training and test set and the training set was further split 75/25 into a development and validation set to search optimal hyperparameters. Hyperparameters were selected using a grid search across three model types, logistic regression, random forests and light gradient boosted machine models. The model with the highest area-under-the ROC curve (AUROC) after 3-fold cross-validation was chosen to generate the probabilities of being observed and propensity scores for the entire dataset. To avoid the undue influence of extreme propensity scores, we applied symmetric trimming and only included patients with propensity scores between 0.05 and 0.95. We additionally only included patients with follow-up time for the treatment outcome under consideration. After adjusting for the observation probability and propensity for treatment, the average treatment effect (ATE) was estimated as follows,

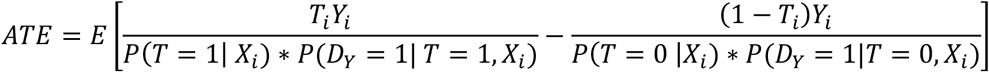

where *i* is the participant, *X* are the covariates, *T* = 1 if given a first-line treatment, *D*_*Y*_ = 1 if the patient was followed for at least the outcome variable’s follow up period and *Y* is the outcome, which is treated as missing when *D*_*Y*_ = 0. Confidence intervals for the propensity scores and *ATE* were generated using bootstrapping^16^. Feature importance for both models was determined using Shapley Additive Explanation values (SHAP values)^17^. Figure 2 represents the analytic pipeline.

**Figure 2:**
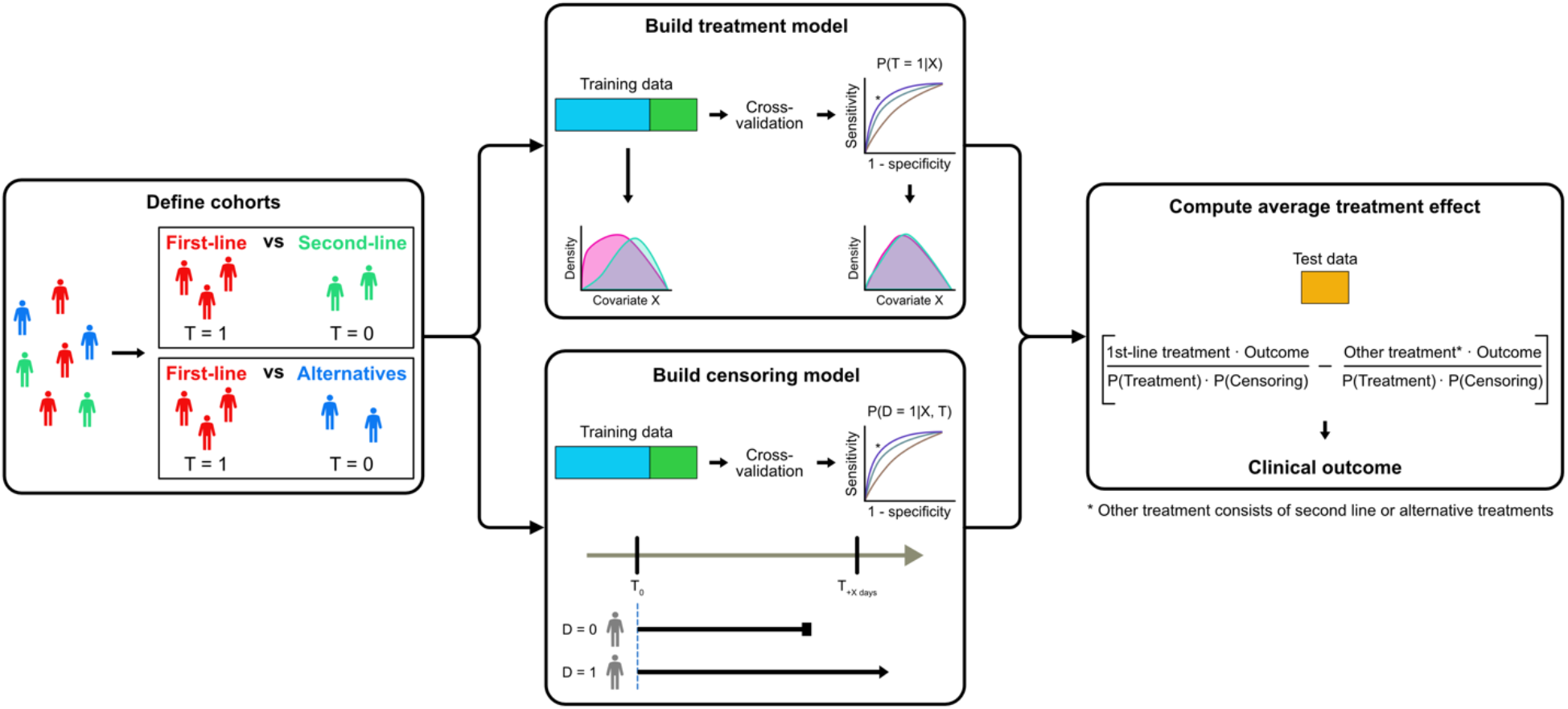
Analytic pipeline. We built and separately analyzed cohorts for first-line versus second-line and first-line versus alternative treatment. Eighty percent of the total data was set aside for training and this was further split 75/25 into development (blue) and validation (green) datasets. Two models were then run to estimate the probability of treatment and of being observed through the outcome period post-diagnosis. We used a 3-fold cross-validation to select the model with the highest AUROC, indicated by the asterisk. Average treatment effect for a given outcome was estimated on test data (yellow) by the risk difference between those receiving first-line treatment or another treatment (second-line or alternative) after normalizing for the probability of receiving a treatment and of being observed at the end of the outcome’s follow-up period (e.g 30 days). Abbreviations, T, treatment, X, covariates, D, observed.

We assessed for residual confounding by assessing treatment effect on three negative control outcomes, fibrocystic disease of the breast, hernia and fracture ^4,18^. These were selected based on domain knowledge and a comprehensive literature search that found no evidence of a causal association with exposure to our antibiotics of interest. Treatment effect was calculated by estimating the prevalence of each negative control outcome at 1 month and 3 months after exposure.

The primary analysis used the model specified by domain expert knowledge. Sensitivity analyses included subgroup analysis in patients who were admitted within a 30-day period after their initial diagnosis of UTI and with the model specified by `OMOP-learn`, using the same analysis pipeline. All analyses were run in Python v3.85 and source code to reproduce all analyses is available at GitHub (https://github.com/clinicalml/uti-causal-inference/).

## RESULTS

### Baseline cohort description

The study flow diagram is shown in Figure 3 and baseline cohort characteristics are summarized in Table 1.

**Figure 3:**
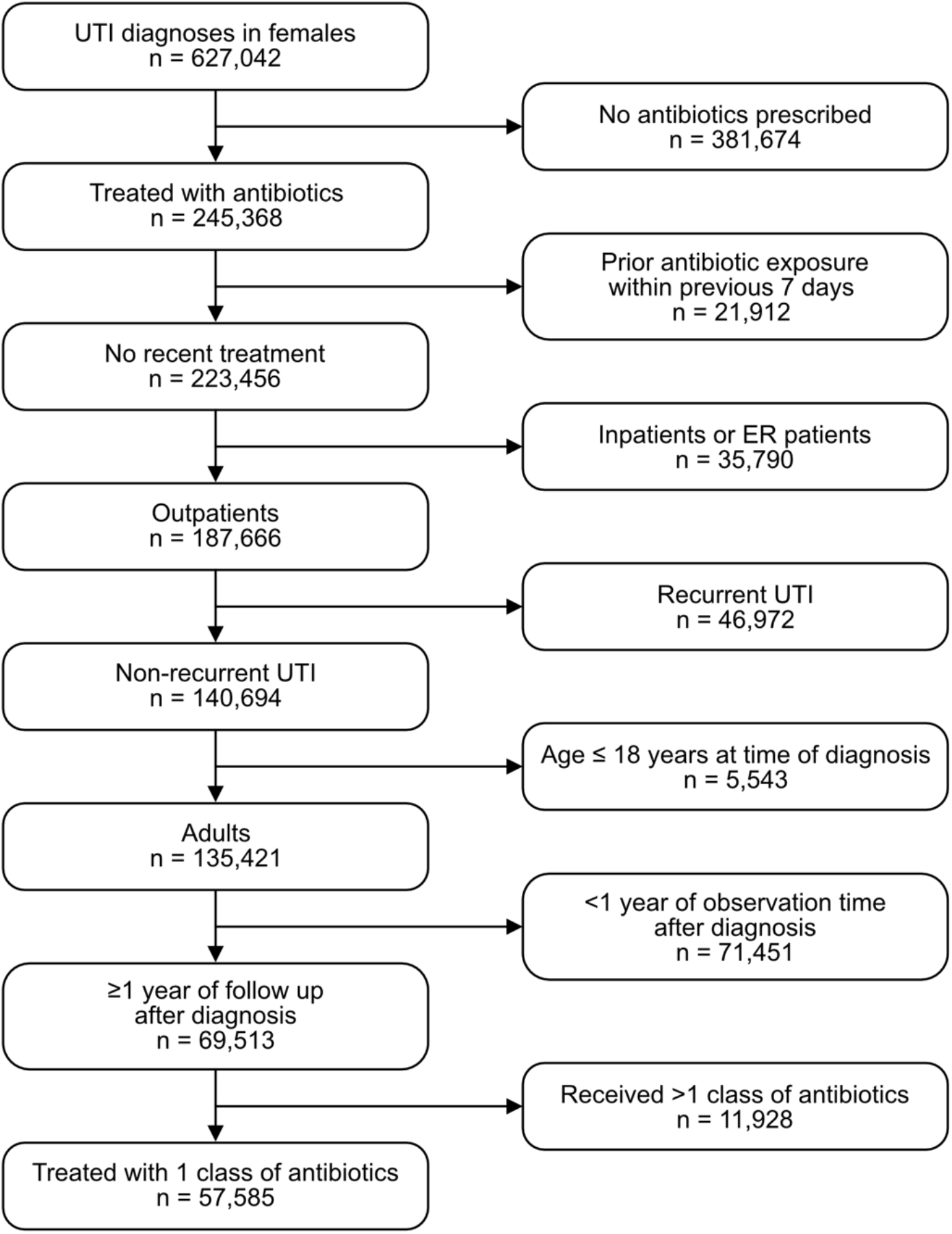
Study flow diagram. Sample sizes indicate UTI diagnoses.

**Table 1.** Baseline characteristics of cohort. Unless otherwise indicated, values represent sample size and column percentage. * p <0.05 compared to first-line cohort

| Baseline characteristics | Full cohort | First line | Second line | Alternative |
| --- | --- | --- | --- | --- |
| Sample size (UTI diagnoses) | 57,585 | 35,018 | 21,140 | 1,427 |
| Age (median, IQR) | 52 (29) | 49 (30) | 57 (29)* | 59 (35)* |
| Fever at presentation | 649 (1.1) | 207 (0.6) | 390 (1.8)* | 52 (3.6)* |
| Urinalysis ordered at presentation | 13,713 (23.8) | 7,365 (21.0) | 5,937 (28.1)* | 411 (28.8)* |
| Blood test ordered at presentation | 2,248 (3.9) | 1,040 (3.0) | 1,057 (5.0)* | 151 (10.6)* |
| Menopause | 3,852 (6.7) | 2,320 (6.6) | 1,424 (6.7) | 108 (7.6) |
| UTI in past year | 5,863 (10.2) | 3,418 (9.8) | 2,257 (10.7)* | 188 (13.2)* |
| Underlying conditions |  |  |  |  |
| Hypertension | 21,026 (36.5) | 10,742 (30.7) | 9,525 (45.1)* | 759 (53.2)* |
| Diabetes Mellitus | 8,298 (14.4) | 4,056 (11.6) | 3,910 (18.5)* | 332 (23.3)* |
| Arthritis | 11,220 (19.5) | 6,122 (17.5) | 4,698 (22.2)* | 400 (28.0)* |
| Cancer | 5,684 (9.9) | 2,847 (8.1) | 2,639 (12.5)* | 198 (13.9)* |
| Chronic kidney disease | 2,992 (5.2) | 1,364 (3.9) | 1,458 (6.9)* | 170 (11.9)* |
| Autoimmune | 2,956 (5.1) | 1,639 (4.7) | 1,207 (5.7)* | 110 (7.7)* |
| Thyroid Disorder | 292 (0.5) | 148 (0.4) | 136 (0.6) | 8 (0.6) |
| Year of UTI episode |  |  |  |  |
| 2012-2014 | 7,376 (12.8) | 3,435 (9.8) | 3,792 (17.9)* | 149 (10.4) |
| 2015-2017 | 26,770 (46.5) | 14,757 (42.1) | 11,420 (54.0)* | 593 (41.6) |
| 2018-2021 | 23,439 (40.7) | 16,826 (48.1) | 5,928 (28.0)* | 685 (48.0) |
| Provider specialties |  |  |  |  |
| Family medicine | 17,523 (30.4) | 9,710 (27.7) | 7,426 (35.1)* | 387 (27.1) |
| Internal medicine | 8,025 (13.9) | 3,858 (11.0) | 3,930 (18.6)* | 237 (16.6)* |
| Emergency care | 4,757 (8.3) | 2,968 (8.5) | 1,689 (8.0)* | 100 (7.0) |
| Obstetrics / Gynecologist | 2,839 (4.9) | 2,124 (6.1) | 684 (3.2)* | 31 (2.2)* |
| Other non-urology specialist | 6,729 (11.7) | 5,119 (14.6) | 1,480 (7.0)* | 130 (9.1)* |
| Urology | 1,498 (2.6) | 907 (2.6) | 522 (2.5) | 69 (4.8)* |
| Others | 2,385 (4.1) | 1,433 (4.1) | 818 (3.9) | 134 (9.4)* |

The final analysis cohort consisted of 57,585 episodes of UTI occurring in 49,037 patients. Of these, first-line antibiotics were prescribed in 35,018 (61%) episodes, second-line antibiotics were prescribed in 21,140 (37%) episodes and alternative antibiotics were prescribed in 1,427 (2%) episodes. Compared to those prescribed with first-line antibiotics, patients prescribed second-line antibiotics were older, more likely to present with fever, more likely to have laboratory tests ordered, and had a higher comorbidity burden. Those prescribed alternative treatments were similarly older, more likely to be febrile and had a higher comorbidity burden. They were also more likely to be seen in the emergency room.

### Primary outcomes

For the domain expert-derived features, a light gradient boosting machine was the best model for predicting censorship as well as the likelihood of treatment (details of performance in Supplementary Results and Supplementary Figures). The top 5 covariates predicting first-line versus second-line therapy were year at UTI diagnosis, patient age, whether the provider was an advanced specialist, internal medicine or family medicine doctor. The top 5 covariates predicting first-line versus alternative therapy were age, whether the provider was an obstetrics / gynecologist, and whether the individual had received beta-lactams in the previous 2 years.

Patients with UTI who were prescribed first-line antibiotics had a lower probability of an inpatient and outpatient revisit within 30 days compared to those who received a second-line antibiotic (adjusted risk difference = -1.8% [95% CI -2.4% to -1.1%]). Relative to alternative beta-lactam treatments, patients prescribed first-line antibiotics for UTI had a 6.4% [95% CI -10.1% to -3.2%] lower probability of inpatient or outpatient revisit at 30 days (Figure 4). For both comparisons, these findings were driven largely by individuals with uncomplicated UTI, but were also observed in those with pyelonephritis or sepsis to a lesser extent.

**Figure 4.**
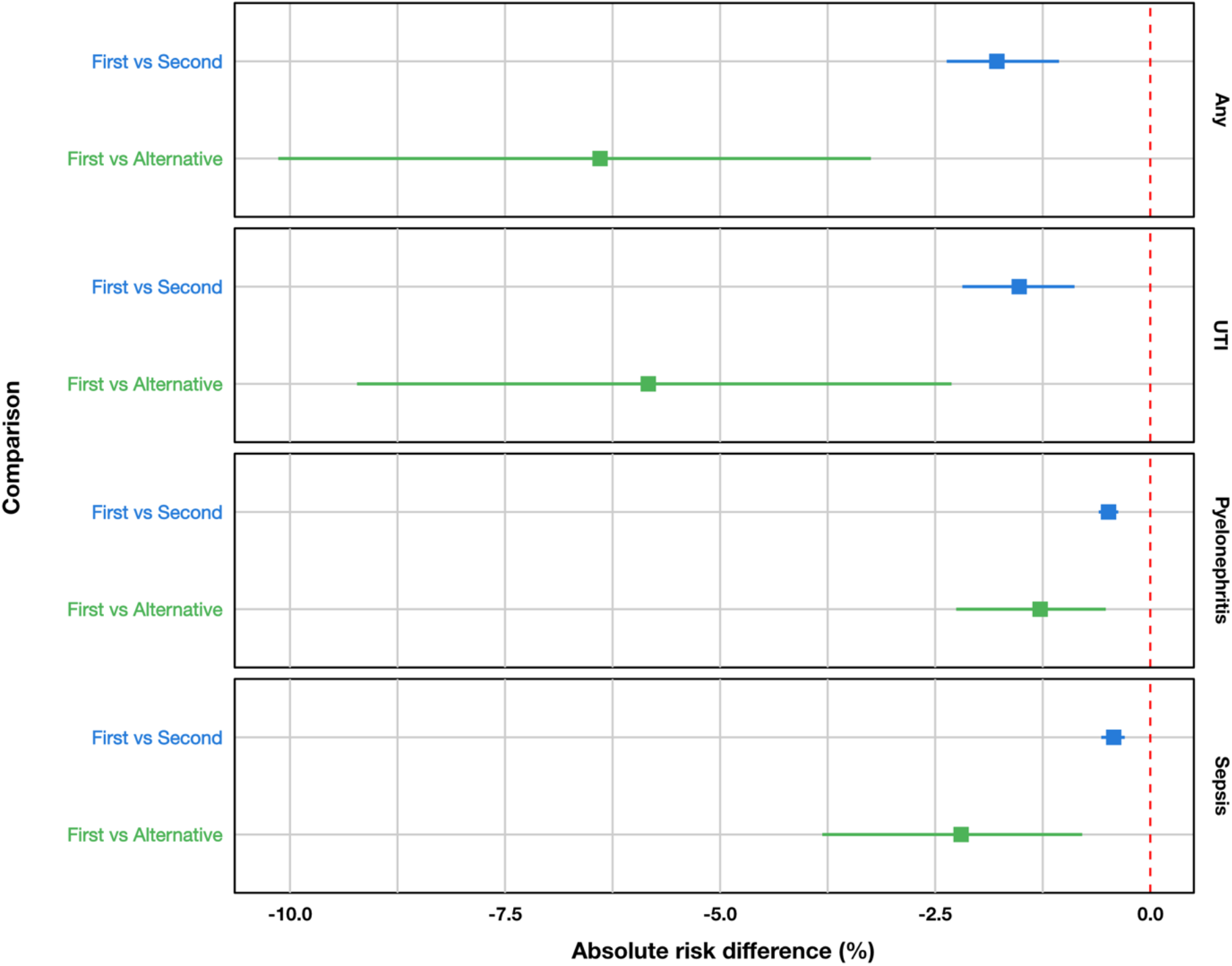
Adjusted rate difference for revisits for patients receiving first-line versus second-line antibiotics, and first-line versus alternative treatments, after adjusting for potential confounding factors and censoring.

### Secondary outcomes

In terms of adverse events, receipt of first-line antibiotics was associated with a slightly increased risk for skin-related adverse events (adjusted risk difference +0.4% [95% CI: +0.2% to +0.5%]) compared to second-line antibiotics and a decreased risk of acute kidney injury within 30 days of treatment (adjusted risk difference -0.3% [95% CI: -0.5% to -0.2%]) (Figure 5). There was no difference in the risk for *C. difficile* infection between the treatment groups.

**Figure 5.**
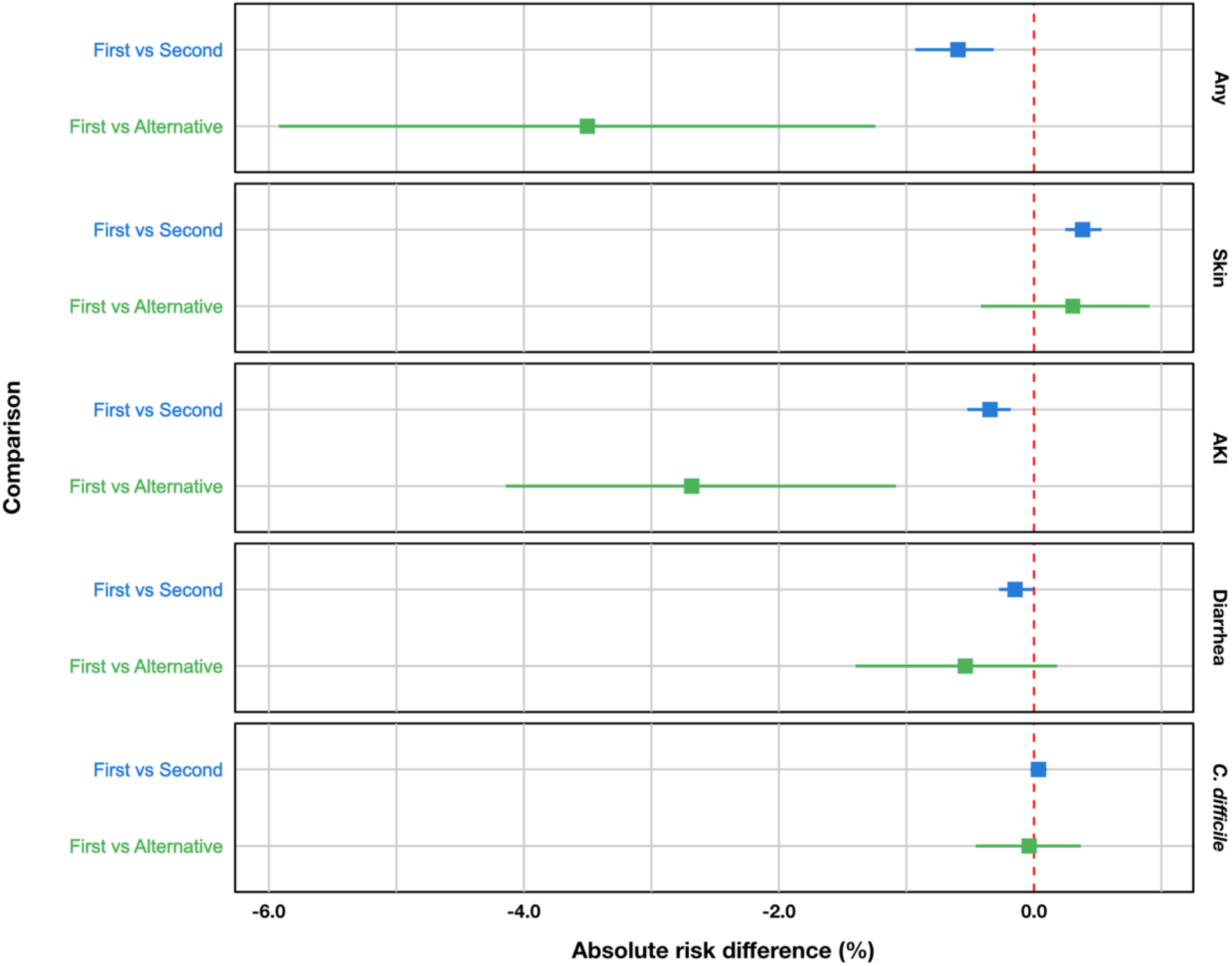
Adjusted rate difference for treatment-related adverse effects for patients receiving first-line versus second-line antibiotics, and first-line versus alternative treatments, after adjusting for potential confounding factors and censoring.

Receipt of first-line antibiotics for UTI was associated with a lower risk of acute kidney injury at 30 days (adjusted risk difference -2.7% [95% CI: -4.1% to -1.1%]), relative to receiving an alternative treatment (Figure 5). There was no difference in the risk of skin-related adverse events, diarrhea, and *C. difficile* infection at 90 days.

### Negative Control Outcomes and Sensitivity Analyses

There were no differences in the 1-month and 3-month risk for the three negative control outcomes regardless of whether the patient received a first-line, second-line or alternative treatment (Figure S3). We observed little to no difference between treatment arms in the sensitivity analysis for patients who had an inpatient revisit within 30 days (1^st^ line versus 2^nd^ line -0.1%, [95% CI: -0.3% to 0.0%]; 1^st^ line versus alternative 0.3%, [95% CI -0.7% to 1.1%], Figures S4). Additionally, results obtained from the model specified by `OMOP-learn` were similar across all comparators and outcomes to the domain expert specified model (Supplementary Results and Figures S5, S6 and S7). For the `OMOP-learn` model, first-line antibiotics had better efficacy than second-line antibiotics as measured by lower risk of medical revisits (−2.1% [95% CI: -2.9% to - 1.6%]) overall, and in those with inpatient revisits (−0.2% [95% CI: -0.3% to 0.0%]). They also had a lower overall risk of any adverse events (−0.7% [95% CI: -1.0% to -0.4%]) but a higher rate of skin-related adverse events (adjusted risk difference: 0.3% [95% CI: 0.2% to 0.4%]). Similar results were observed in the comparison between first-line antibiotics and alternative antibiotics using the `OMOP-learn` derived model.

## DISCUSSION

Using a large contemporary real-world dataset, we demonstrate that IDSA guidelines for treatment of uncomplicated UTI remain robust in terms of both efficacy and adverse events, despite major changes in the epidemiology of antibiotic resistance^21,22^. Unless a patient has a history of drug resistance, or intolerance or lives in a region where local rates of resistance are high, nitrofurantoin and trimethoprim-sulfamethoxazole remain the treatments of choice. We replicated our domain-expert derived results with an automated feature building package applied to a common data model, thereby supporting the hypothesis that complex causal inference analyses combined with careful cohort selection can be semi-automatable. This will help promote reproducibility of our findings in other health systems and opens inquiry into other important clinical questions.

We observed a small increase in rates of revisits for patients receiving second-line therapy relative to those receiving first-line antibiotics. This result is surprising as fluoroquinolones are thought to be equivalent or superior to nitrofurantoin and TMP-SMX in terms of clinical efficacy^23^. The differences were limited to outpatients with a diagnosis of lower urinary tract infection and were much less pronounced for inpatients, suggesting the benefit of first-line treatments is restricted to classic presentations of uncomplicated UTI. Follow up visits soon after treatment may be driven by drug intolerance, toxicity or by selection of a drug to which an organism is resistant. The latter may be a possible explanation for why people treated with nitrofurantoin and TMP-SMX had fewer revisits. Recent work has suggested that rates of resistance to nitrofurantoin remain low despite its widespread use and may be due to a high barrier to resistance^24^. While resistance to TMP-SMX is more common, clinicians are less likely to use this drug based on IDSA guidance that recommends avoiding it when rates of local resistance exceed 20%, which is a common scenario throughout the United States. In contrast, resistance to fluoroquinolones is most often mediated by the accumulation of mutations in a single gene often in response to antibiotic exposure. Given the high rate of fluoroquinolone prescription in the community, this may increase the risk for prescribing an agent to which the agent is resistant. This is further complicated by the fact that uncomplicated UTI, is often managed over telephone and without culture data. Lastly, given that prescribers are prone to prescribe the same antibiotic^25–27^, the impact of prior exposure may be more likely to lead to selection of resistance if that antibiotic is a fluoroquinolone and the patients are otherwise healthy outpatients with a low risk for colonization by drug-resistant organisms.

We applied two approaches to feature construction to correct for confounding. Domain expert-derived features are derived from expert knowledge on the biologic mechanisms of disease and real-world experience with managing uncomplicated UTI. These features have the advantage of theoretical backup from established pathophysiology and clinical data, but suffer from the possibility of missing potential confounders, especially when the disease has diverse mechanistic pathways or is not well-understood. In contrast, `OMOP-learn`^14^, which captures all information available in the data without prior knowledge of its relationship with the disease, lowers the probability of missing confounders but comes at the expense of including a large number of non-relevant covariates. Our study provides an empirical demonstration that extracting features under the `OMOP-learn` framework can yield conclusions comparable to that domain expert-derived features, which supports application of causal inference methods using automatic feature generation in the medical context.

Recent work has shown that carefully constructed retrospective cohorts with proper statistical adjustment can provide robust results that complement findings from prospective randomized controlled trials^28^. However, as with all observational studies, there is a possibility that our results may be biased due to residual confounding. We believe the degree of confounding is small as we adjusted for both covariate-dependent censoring and treatment indication, which are the major forms of confounding we expect to impact our results. This is further supported by the results of the negative control outcome analysis, which shows an equal distribution of control outcomes between treatment arms. The consistency in the strength and direction of our outcomes between domain-expert derived and `OMOP-learn` derived features lends further strength to the validity of our findings.The major strength of this study is the inclusion of a real-world dataset with a comprehensive collection of covariates translated into a common data model. The rich set of features permits construction of models that better specify causal mechanisms and the use of a common data model enhances the study’s reproducibility for other patient populations. Lastly, large observational datasets offer the opportunity to gain real-world insight that is both up to date and representative of the patients presenting with disease in practice today.

Other limitations of our study are that the prevalence of certain comorbidities is lower in our cohort than in the general population. This may partly reflect the limited scope of our data, which comes from a single health insurer primarily based in Southeast Pennsylvania but may also reflect our inclusion criteria, which intentionally restricted our analyses to people with uncomplicated UTI. We also had limited data on patient race, ethnicity and socioeconomic status, which precluded our ability to assess for fairness across diverse subpopulations. Future work should seek to reproduce our analysis using larger datasets with more diverse populations to ensure equity. The increase in prescription of first-line antibiotics over time, which likely reflects the effect of guideline dissemination and promotion of antibiotic stewardship^29–31^, should not by itself bias outcomes, assuming care practices did not dramatically change over the study period.

In conclusion, our results provide reassurance that guideline-concordant therapy remains the optimal treatment decision for uncomplicated UTI. The application of an automated feature extraction package for datasets translated into a common data model, combined with a rigorous analytic pipeline, is a promising approach to assess the impact of guideline-directed therapy in real-world populations and over time.

## Data Availability

All data produced in the present study are available upon reasonable request to the authors

