## Supplemental_materials for "Reassessing the management of uncomplicated urinary tract infection: A retrospective analysis using machine learning causal inference"

**SUPPLEMENT**

**Table of contents**


| ID | Description | Page |
|---|---|---|
| Supplementary results | | |
| 1 | Performance of propensity score and confounding models | S2 |
| Supplementary tables | | |
| 1 | List of ICD-10-CM diagnosis codes defining diagnosis of UTI | S3 |
| 2 | List of procedures and diagnoses codes for exclusion from the cohort | S4 |
| 3 | List of conditions included in adverse event categories | S5 |
| 4 | List of conditions and medications included as windowed history features in domain expert-derived features | S6 |
| 5 | Prior antibiotic exposures for treatment groups | S7 |
| Supplementary figures | | |
| S1 | Calibration plot for propensity models | S8 |
| S2 | Shapley value plots for feature importance | S9 |
| S4 | Adjusted rate differences in negative control outcomes between $1^{st}$ vs $2^{nd}$ and $1^{st}$ vs alternative treatments | S10 |
| S3 | Adjusted rate differences in treatment efficacy between $1^{st}$ vs $2^{nd}$ and $1^{st}$ vs alternative treatments for inpatients only | S11 |
| S5 | Comparison of domain expert derived vs OMOP features for treatment efficacy, stratified by $1^{st}$ vs $2^{nd}$ and $1^{st}$ vs alternative treatments | S12 |
| S6 | Comparison of domain expert derived vs OMOP features for adverse effects, stratified by $1^{st}$ vs $2^{nd}$ and $1^{st}$ vs alternative treatments | S13 |
| S7 | Comparison of domain expert derived vs OMOP features for negative control outcomes, stratified by $1^{st}$ vs $2^{nd}$ and $1^{st}$ vs alternative treatments | S14 |

### SUPPLEMENTARY RESULTS

Performance of propensity score and confounding models

First-line versus second-line antibiotics

For the domain expert-derived features, the best model predicting censorship for 15 and 90-day loss-to-follow up was a light gradient boosting machine, with AUROCs of 0.68, 0.66. The best censorship model for 30-day loss-to-follow up was a logistic regression with AUROC of 0.66. For predicting receipt of first-line or second-line antibiotics (to assess confounding by indication), the best model was also a light gradient boosting machine, with an AUROC of 0.73. Calibration curves are shown in Supplementary Figure 1a. Using SHAP values, the top 5 covariates contributing to the prediction include year at UTI diagnosis, patient age, and provider specialties (whether the provider was an advanced specialist, internal medicine or family medicine doctor).

First-line versus alternative antibiotics

For the domain expert-derived features, the best model predicting censorship for 30 and 90-day loss-to-follow up was a light gradient boosting machine, with AUROC of 0.62 and 0.60. The best censorship model for 15-day loss-to-follow up was a random forest with AUROC of 0.64. For predicting first-line or alternative antibiotics, the best model was a logistic regression, with AUROC 0.70 and the calibration curve is shown in Supplementary Figure 1b. Using SHAP values, the top 5 covariates contributing to the prediction include patient age, physician specialty (Ob / Gyn), and prior receipt of alternative antibiotics in the prior 24 months.

| Diagnosis description | ICD-10-CM code |
| ---: | --- |
| Urinary tract infectious disease | N39.0 |
| Acute cystitis | N30.00 |
| Cystitis | N30.90 |
| Hematuria co-occurrent and due to cystitis | N30.01 |
| Hematuria co-occurrent and due to acute cystitis | N30.01 |

**Supplementary Table 1. List of ICD-10-CM diagnosis codes defining diagnosis of urinary tract infection.**

| Excluded if occurred within 180 days before UTI diagnosis | |
|---|---|
| Pregnancy | Z33.1 |
| Pyelonephritis | N12 |
| Urinary catheterization | Z96.0 |
| Procedures related to central venous catheter | Z95.828 |
| Any surgery or mechanical ventilation | Z98.890, Z99.11 |
| Hemodialysis | Z49.31 |
| Parenteral nutrition | 3E0336Z |

| Excluded if ever occurred before UTI diagnosis | |
|---|---|
| Neurogenic bladder | N31.9 |
| Spina bifida | Q05.9 |
| Malignancies of the urinary tract | C68.8 |
| Gynecological Malignancies | C55 |

**Supplementary Table 2. List of procedures (ICD-10-PCS) and diagnoses (ICD-10-CM) codes for exclusion from the cohort.**

| | |
|---|---|
| **C. *difficile* infection** | |
| *Clostridium difficile* colitis, *Clostridioides difficile* infection | A04.72 |
| **Skin** | |
| Dermatitis due to drug AND/OR medicine taken internally | L27.0 |
| Pruritic rash | L28.2 |
| Urticaria | L50.9 |
| Eruption | R21 |
| Contact dermatitis due to drugs AND/OR medicine | L23.3 |
| **Gastrointestinal** | |
| Diarrhea | R19.7 |
| **Acute kidney injury** | |
| Acute renal failure syndrome | N17.9 |

**Supplementary Table 3. List of conditions included in adverse event categories.**

| Diagnosis description | ICD-10-CM code |
|---|---|
| Addison’s disease | E27.1 |
| Arthritis | M19.90 |
| Catheter-related conditions | T80.219S |
| Celiac disease | K90.0 |
| Chronic kidney disease | N18.9 |
| Corticosteroid | T38.0X1A |
| Dermatomyositis | M33.90 |
| Diabetes mellitus | E11.9 |
| Graves disease | E05.00 |
| Hashimoto thyroiditis | E06.3 |
| Hemodialysis | Z99.2 |
| HIV | Z21 |
| Hypertension | I10 |
| Lupus erythematosus | L93.0 |
| Malignancies | C80.1 |
| Menopause | Z78.0 |
| Morbid obesity | E66.01 |
| Multiple sclerosis | G35 |
| Myasthenia gravis | G70.00 |
| Pernicious anemia | D51.0 |
| Reactive arthritis | M02.30 |
| Rheumatoid arthritis | M06.9 |
| Sjogren’s disease | M35.00 |
| Transplantation | Z94.9 |
| Urinary incontinence | R32 |

**Supplementary Table 4. List of conditions and medications included as windowed history features in domain expert-derived features.**

| Treatment group | n | NIT | SXT | CIP | OFX | LVX | AMC | CPD | CFR | Any |
|---|---|---|---|---|---|---|---|---|---|---|
| All patients | 57,585 | 4.5 | 4.6 | 5.5 | 0.5 | 2.2 | 5.2 | 0.0 | 0.2 | 19.3 |
| First-line | 35,018 | 5.2 | 4.8 | 4.1 | 0.5 | 1.6 | 5.1 | 0.0 | 0.2 | 18.3 |
| Second-line | 21,140 | 3.3 | 4.2 | 7.6 | 0.4 | 3.2 | 5.0 | 0.0 | 0.1 | 20.4 |
| Alternatives | 1,427 | 5.5 | 4.8 | 6.6 | 0.6 | 3.4 | 11.0 | 0.2 | 0.5 | 26.6 |
| NIT | 20,064 | 6.3 | 3.7 | 4.3 | 0.5 | 1.4 | 5.2 | 0.0 | 0.2 | 18.2 |
| SXT | 14,954 | 3.7 | 6.3 | 4.0 | 0.4 | 1.8 | 5.0 | 0.0 | 0.2 | 18.4 |
| CIP | 18,593 | 3.2 | 4.1 | 7.7 | 0.4 | 2.5 | 5.0 | 0.0 | 0.1 | 19.8 |
| OFX | 17 | 11.8 | 0.0 | 11.8 | 0.0 | 0.0 | 11.8 | 0.0 | 0.0 | 29.4 |
| LVX | 2,530 | 4.1 | 4.9 | 7.3 | 0.5 | 8.3 | 4.9 | 0.1 | 0.2 | 24.5 |
| AMC | 1,235 | 5.8 | 5.0 | 6.5 | 0.7 | 3.5 | 12.2 | 0.0 | 0.1 | 27.5 |
| CPD | 124 | 1.6 | 4.0 | 7.3 | 0.0 | 3.2 | 3.2 | 2.4 | 0.0 | 16.9 |
| CFR | 68 | 8.8 | 2.9 | 7.4 | 0.0 | 2.9 | 2.9 | 0.0 | 8.8 | 27.9 |

**Supplementary Table 5. Prior antibiotic exposures for treatment groups.** Exposure represents the 6-month history for patients in the indicated treatment group.

Abbreviations

- NIT, nitrofurantoin
- SXT, trimethoprim-sulfamethoxazole
- CIP, ciprofloxacin
- OFX, ofloxacin
- LVX, levofloxacin
- AMC, amoxicillin-clavulanate
- CPD, cefpodoxime
- CFR, cefadroxil

**A** **B**

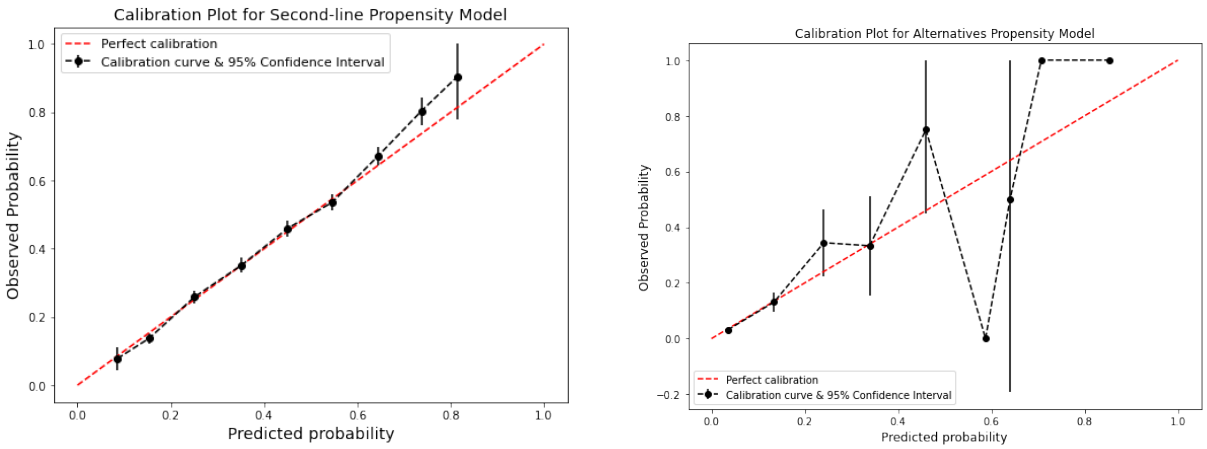


**Supplementary Figure S1.** Calibration plots for propensity for treatment with (A) second-line antibiotics versus first-line antibiotics and (B) alternative antibiotics versus first-line antibiotics. The calibration probability was calculated using validation data comprised of 20% of the dataset.

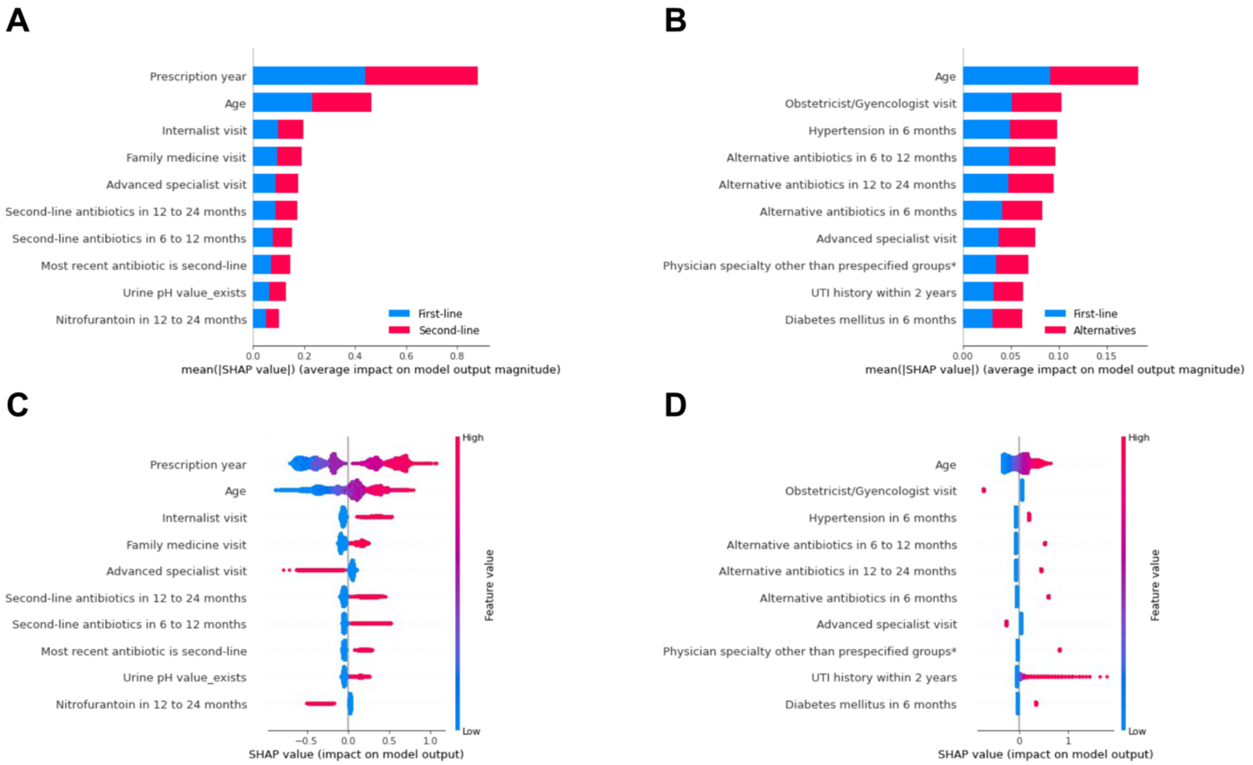


**Supplementary Figure S2.** Shapley value plots showing features ranked by absolute predictive importance for predicting receipt of (A) first-line versus second-line antibiotics and (B) first-line versus alternative antibiotics, and impact of feature level for predicting receipt of (C) first-line versus second-line antibiotics and (D) first-line versus alternative antibiotics.

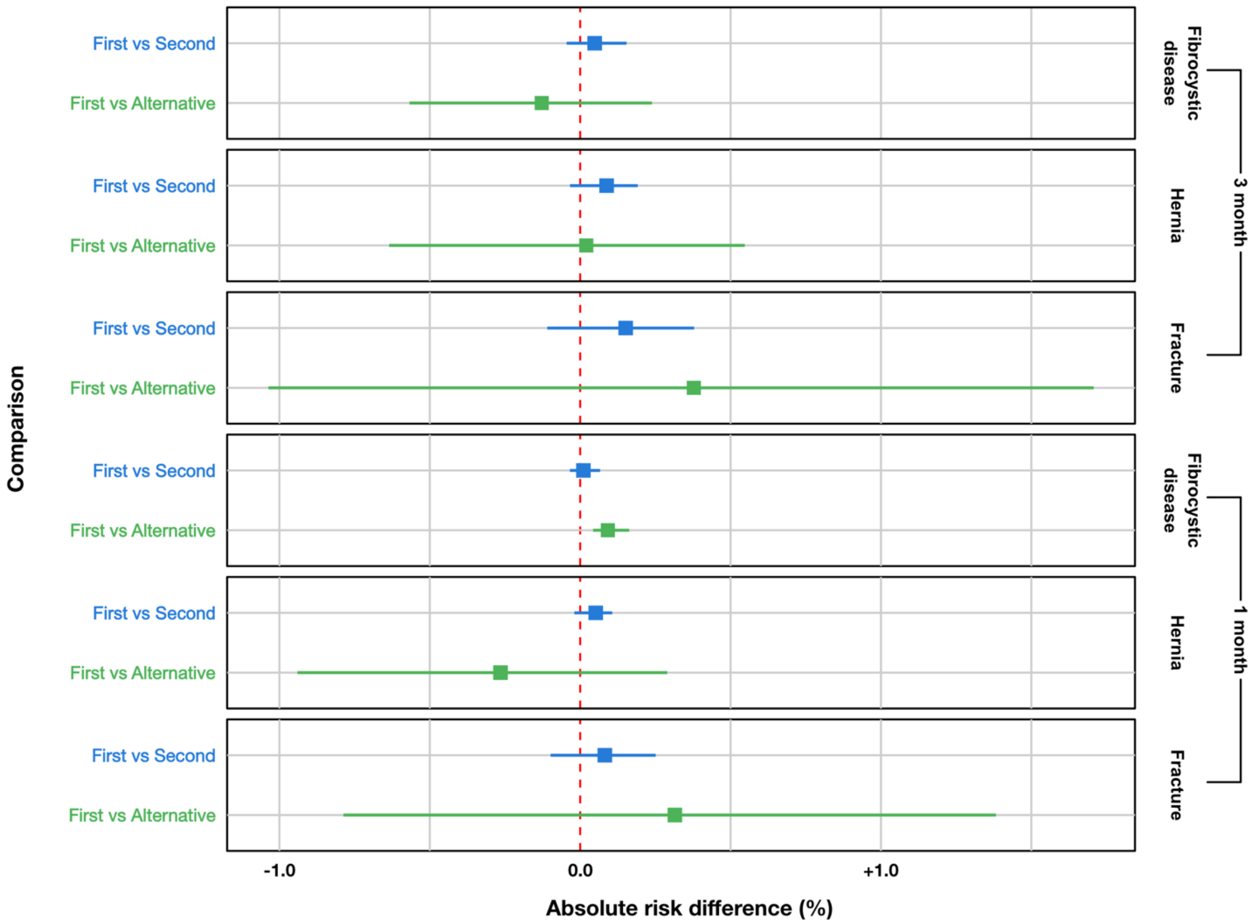


**Supplementary Figure S3.** Adjusted rate difference for negative control outcomes for patients receiving first-line versus second-line antibiotics, and first-line versus alternative treatments, after adjusting for potential confounding factors and censoring.

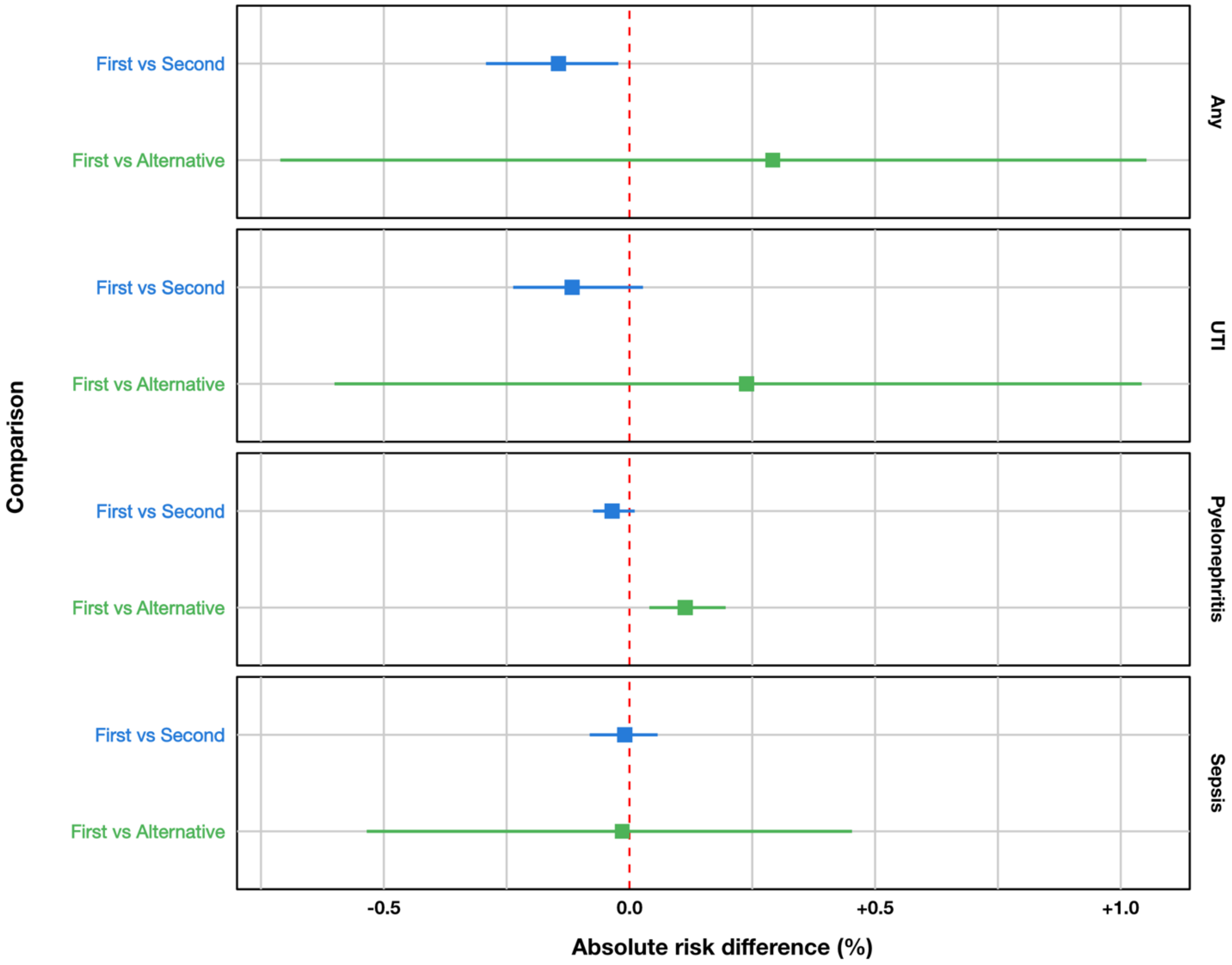


**Supplementary Figure S4.** Adjusted rate difference for revisits resulting in inpatient admission for patients who received first-line versus second-line antibiotics, and first-line versus alternative treatments, after adjusting for potential confounding factors and censoring.

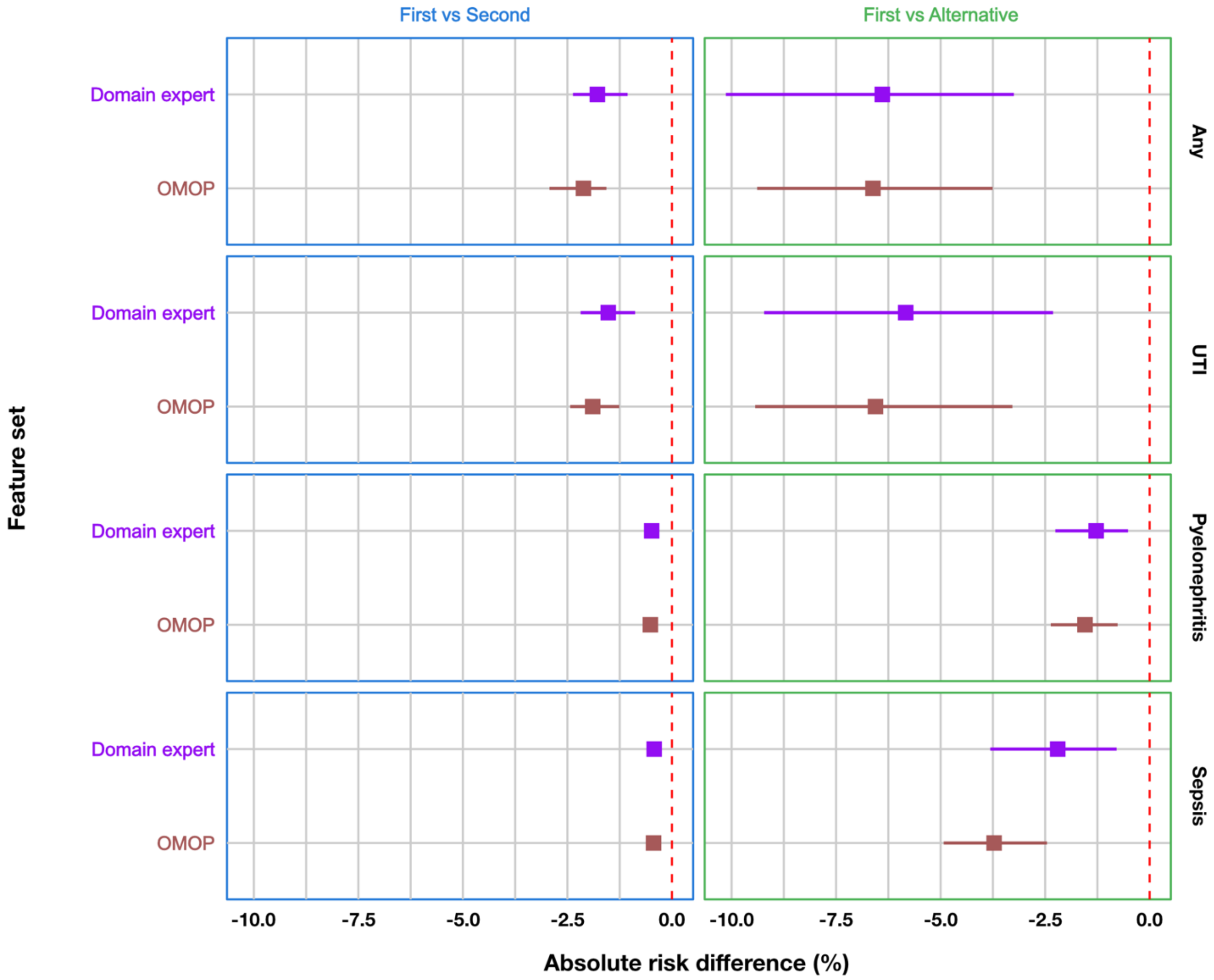


**Supplementary Figure S5.** Comparison of adjusted rate difference for treatment efficacy using domain expert-derived features versus OMOP features, stratified by patients who received first-line versus second-line antibiotics, and first-line versus alternative treatments. Treatment efficacy was estimated by 30-day revisits overall, and for UTI, pyelonephritis and for sepsis.

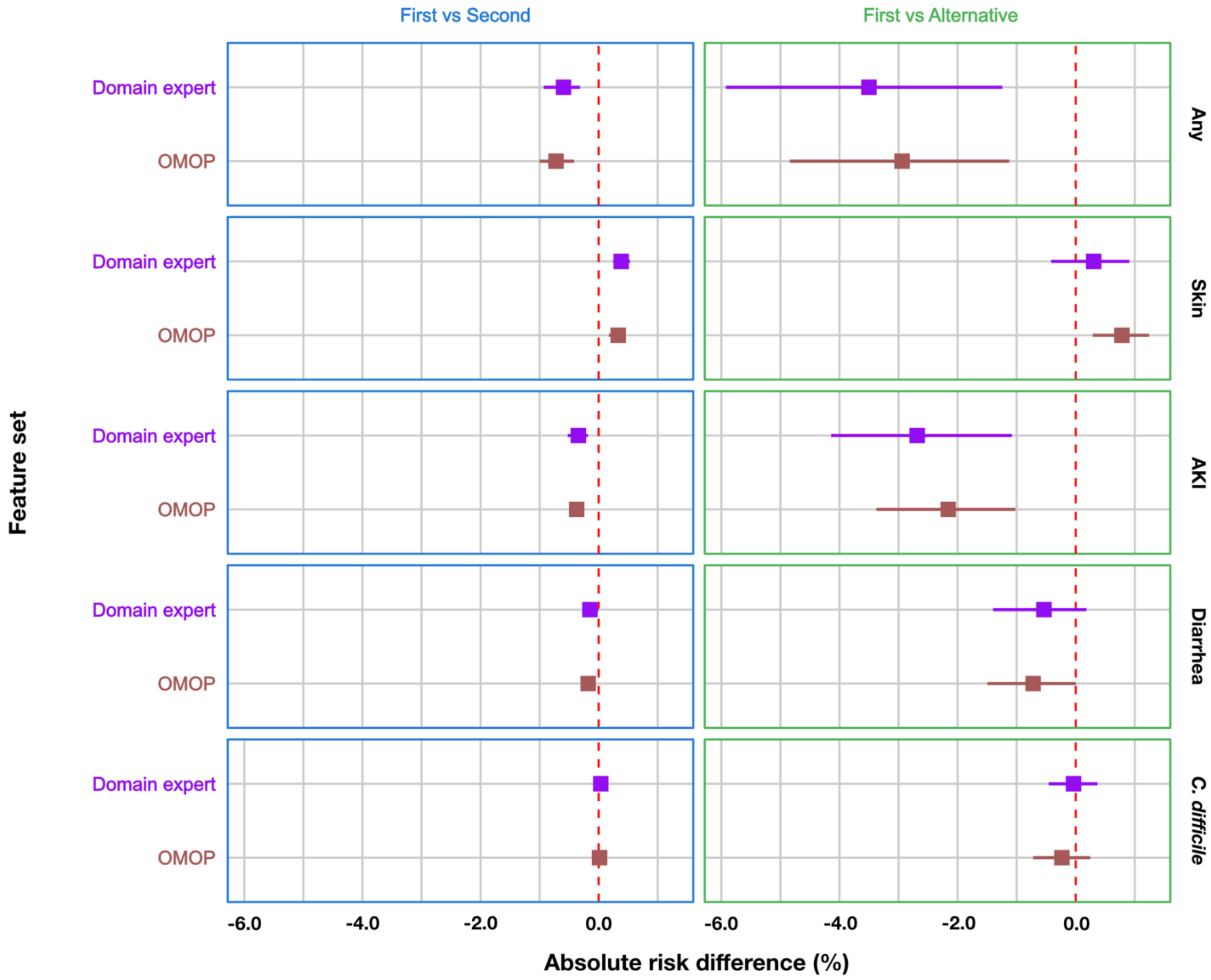


**Supplementary Figure S6.** Comparison of adjusted rate difference for adverse events using domain expert-derived features versus OMOP features, stratified by first-line versus second-line antibiotics, and first-line versus alternative treatments.

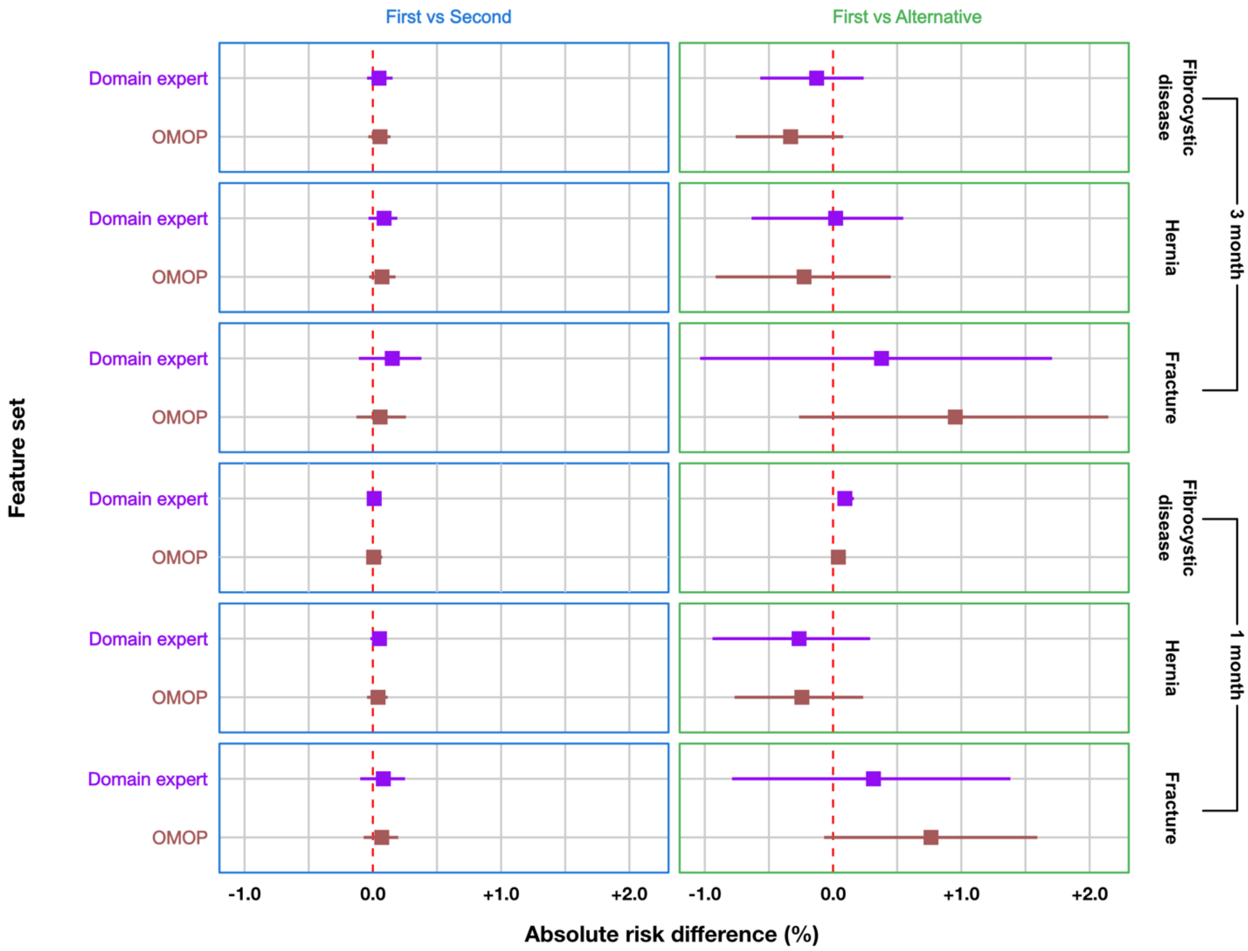


**Supplementary Figure S7.** Comparison of adjusted rate difference for negative control outcomes using domain expert-derived features versus OMOP features, stratified by first-line versus second-line antibiotics, and first-line versus alternative treatments.
